# Comprehensive Surveillance of SARS-CoV-2 Spread Using Wastewater-based Epidemiology Studies

**DOI:** 10.1101/2020.08.18.20177428

**Authors:** Manupati Hemalatha, Uday Kiran, Santosh Kumar Kuncha, Harishankar Kopperi, C. G. Gokulan, S Venkata Mohan, Rakesh K Mishra

**Author notes:** These authors contributed equally to the work (Manupati Hemalatha, Uday Kiran, Santosh Kumar Kuncha).

## Abstract

SARS-CoV-2 pandemic is having a devastating effect on human lives. Individuals who are symptomatic/asymptomatic or have recovered are reported to have/will have serious health complications in the future, which is going to be huge economic burden globally. Given the wide-spread transmission of SARS-CoV-2 it is almost impossible to test each and every individual for the same and isolate them. Recent reports have shown that sewage can be used as a holistic approach to estimate the epidemiology of the virus. Here we have estimated the spread of SARS-CoV-2 in the city of Hyderabad, India which is populated with nearly 10 million people. The sewage samples were collected from all the major sewage treatment plants (STPs) and were processed for detecting the viral genome using the standard RT-PCR method. Based on the average viral particle shedding per individual, the total number of individuals exposed to SARS-CoV-2 (in a window of 35 days) is about 6.6% of the population, which clearly indicates the rate of community transmission and asymptomatic carriers is higher than the number of reported cases. It is important to note here that the samples collected from the inlet of STPs were positive for SARS-CoV-2, while the outlets were negative indicating the efficient treatment of sewage at STPs. These studies are going to be essential to manage the pandemic better and also to assess the effectiveness of control measure.

## 1. Introduction

The surveillance of disease prevalence during pandemic like Corona Virus Disease-19 (COVID-19) is a crucial task considering the spreading rate and high population in different parts of the world. The massive testing of the population to contain the spread of the virus is a challenge to any nation in the present scenario. Moreover, a majority of Severe Acute Respiratory Syndrome Coronavirus-2 (SARS-CoV-2) infected population is asymptomatic. Emerging studies have shown the after effects of COVID-19, which is going to be huge economic burden globally and therefore pressing the importance of not only managing the infected individuals but also to keep the spread to minimum (McKibbin et al., 2020). Considering the present testing capacity and cost incurred, it is impractical to test all the individuals. Thus, there is a need for alternative strategies to assess the disease spread and therefore efficiently allocate resources for disease management.

Even though SARS-CoV-2 is majorly a respiratory pathogen, the persistence and replication of virus in the gastrointestinal (GI) tract and shedding through faeces is established (Wang et al., 2020; Xiao et al., 2020; Zhang et al., 2020; Young et al., 2020; Woelfel et al., 2020). Different independent studies highlighted the presence and replication of SARS-CoV-2 in GI tract and the prolonged shedding of SARS-CoV-2 viral material through faeces during and after active infectious stage (Holshue et al., 2020, Woelfel et al., 2020, Kitajima et al., 2020, Cai et al., 2020, Ling et al., 2020, Wu et al., 2020, La Rosa et al., 2020a; Xiao et al., 2020; Ahmed et al., 2020; Wu et al., 2020; Wurtzer et al., 2020; La Rosa et al., 2020b; Medema et al., 2020).

In this scenario, wastewater-based epidemiology (WBE) studies are suitable to understand and estimate the virus spread in a given population for effective disease surveillance. WBE was earlier used to detect and manage viral diseases such as polio, rotavirus, etc. (Lodder et al., 2012; Lodder et al., 2013; Ahmed et al., 2020). Recent reports employed WBE-based approaches to detect SARS-CoV-2 in sewage water and estimated the percentage of infected individuals in a given population (Ahmed et al., 2020; Wu et al., 2020; Wurtzer et al., 2020; La Rosa et al., 2020b; Medema et al., 2020). The monitoring of SARS-CoV-2 in wastewater could quantify the scale of infection prevailing among the community with a benefit of detecting virus from symptomatic, asymptomatic, or pre-symptomatic cases which manifest as an early-warning sign (Lodder et al., 2020; Medema et al., 2020; Mallapaty et al., 2020). WBE approach will help to minimize the outbreak spread and also serve for future epidemic surveillance (Daughton 2018; Mao et al., 2020).

Here we have studied the spread of SARS-CoV-2 infection in Hyderabad, which is one of the major and densely populated metropolitan cities of India. The wastewater infrastructure of the city was used as an effective resource to access and estimate the spread of SARS-CoV-2 across the city of Hyderabad. Here we have simplified the sample processing method for viral detection by Reverse Transcription-Polymerase Chain Reaction (RT-PCR) and based on the RNA titter the number of infected individuals were estimated.

## 2. Materials and Methods

### 2.1. Sampling sites

The detection of SARS-CoV-2 genetic material in domestic sewage was performed by collecting samples from different sewage treatment plants (STPs) in Hyderabad Metropolitan City, India. Hyderabad (17.37°N 78.48°E) is fifth-largest urban economy in India and is a capital for Telangana state occupies ∼625 square kilometers. It is the fourth-most populous city in India with 10 million residents in the metropolitan region. The raw sewage samples were collected from inlet and outlet points from the STPs with a total coverage of 712 million litres per day (MLD), that receive wastewater from all parts of the city (80% coverage of the existing STPs).

### 2.2 Sampling procedure

The sewage samples were collected from 7^th^ July 2020 to 8^th^ August 2020 taking all the safety measures as per the standard operating procedure (SOP) designed for this purpose (Supplementary Table 1). Sampling was performed in the day time, on the days with no report of rainfall during last 24 hours. A total of 30 samples were collected from 14 inlets and 14 treated outlets of 10 STPs and 2 samples from a gated community. A 10 MLD STP was selected for a time course study to understand the weekly variation in the viral load.

**Table 1:** SARS-CoV-2 RNA C_T_ values (and standard error of mean) detected by real-time RT-PCR of raw sewage samples collected from 10 MLD STP with varied sodium hypochlorite concentrations. n.d. = not detected; PC = Positive Control; NTC = No Template Control.

| Sodium Hypochlorite (%) | $C_T$ Values | | |
| --- | --- | --- | --- |
|  | E-gene | N-gene | ORF1ab |
| 0.1% | 26.58±0.031 | 25.69±0.088 | 26.17±0.098 |
| 0.5% | 27.13 | 26.16±0.120 | 26.970 |
| 1% | 27.46±0.134 | 26.75±0.130 | 26.96±0.159 |
| 2% | n.d. | n.d. | n.d. |
| 3% | n.d. | n.d. | n.d. |
| 4% | n.d. | n.d. | n.d. |
| PC | 25.94 | 25.47 | 23.13 |
| NTC | n.d. | n.d. | n.d. |

**Figure 1:**
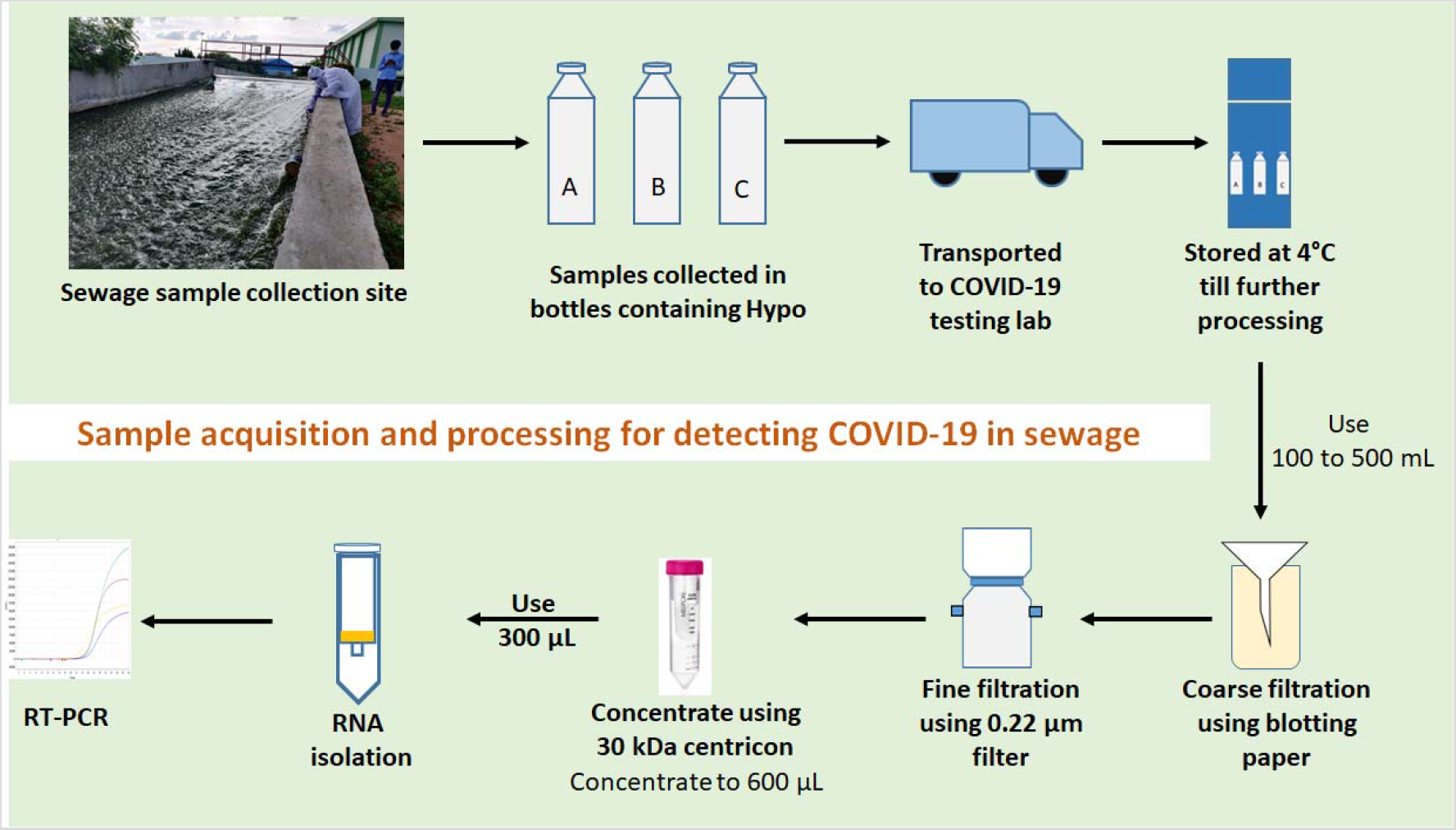
Schematic of sample collection and processing: The sewage samples are collected from STPs in bottles containing Hypo (0.1-4%) followed by shipping to the COVID-19 testing laboratory and stored at 4 °C until further processing. The samples are initially filtered using blotting paper followed by 0.22 μm filter. These filtered samples are further concentrated using 30 kDa centricon filters. The concentrates are used for RNA isolation, the isolated RNA is subjected to RT-PCR using COVID-19 specific primers and probes.

### 2.3 Optimization of disinfectant concentration

Optimum concentration of sodium hypochlorite (5%; Qualigens) addition during sampling was optimized by collecting samples in various initial concentrations of hypochlorite (0.1%, 0.5%, 1%, 2%, 3% and 4%) using samples collected from 10 MLD STP. Grab samples were collected from the inlet point of 10 MLD STP in disposable plastic bottles containing 20 ml of the above mentioned concentrations of sodium hypochlorite. Collected samples were sealed to avoid leakage and wrapped in plastic covers in two layers and transferred to the laboratory immediately and stored at 4^°^C. Samples were processed within 24 h of sampling for the detection of SARS-CoV-2 RNA.

### 2.4. Processing of Samples

Collected samples were subjected to gravity filtration with 1 mm thick blotting sheets to remove the larger particles followed by filtration using 0.2 µm filtration units (Nalgene® vacuum filtration system; Thermo Fisher Scientific, USA) to remove bacteria and other debris. The filtrate was collected in 1000 mL sterile wide-mouth bottles (Borosil, India). 100 mL of the total filtrate was concentrated to ∼600 µl using 30 kDa Amicon® Ultra-15 (Merck Millipore, USA) by centrifuging at 4000 rpm at 4^°^C. The concentrated samples were further processed for RNA isolation. Sample filtration, concentration and processing till detection were performed in Biosafety level 2 (BSL-2). All the materials after use were discarded in biosafety bags followed by decontamination.

### 2.5. RNA extraction and RT-PCR

The RNA extraction from 300 µl of concentrate was performed using QIAamp Viral RNA isolation kit (Qiagen, Germany) by following manufacturer’s protocol. The isolated RNA samples were tested for presence of SARS-CoV-2 RNA using Fosun COVID-19 RT-PCR Detection Kit (Shanghai Fosun Long March Medical Science Co., Ltd, China). The kit detects Envelope (E; ROX labelled), Nucleocapsid (N; JOE labelled) and open reading frame1ab (ORF1ab; FAM labelled) genes of SARS-CoV-2 and the RT-PCR was performed as per manufacturer recommendation. A C_T_ value of 32 corresponds to 300 copies per mL when using this kit (https://www.fda.gov/media/137120/download). The samples were tested either in duplicates or triplicates.

### 2.6. Calculation of number of infected people in a population

We followed two different methods to calculate the number of infected individuals in a given population based on the average number of RNA copies present in the sewage water.

Method: 1 (Ahmed et al., 2020)

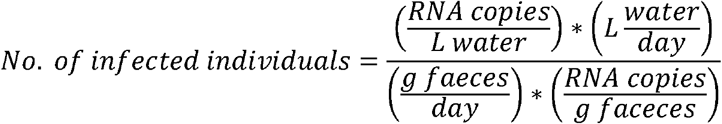

Faeces excreted/person/day = 128 g (Rose et al., 2015)

One positive person sheds 10^7^ RNA copies/g of faeces (maximum estimate, Foladori et al., 2020)

Method 2: (Hellmér et al., 2014)

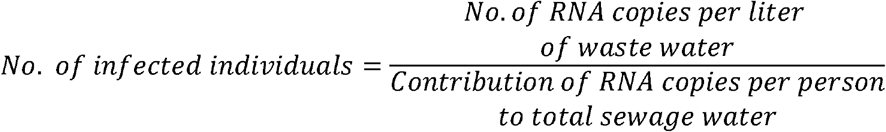

Number of RNA copies excreted per mL of faeces = 10^7^

Volume of faeces excreted = 120 mL (calculated by considering the density of human faeces is 1.07g/mL, (Foladori et al., 2020)). Total RNA copies excreted per person = 1.2×10^9^ copies.

## 3. Results and Discussion

### 3.1 Optimization of Disinfection Concentration

Sodium hypochlorite was used to disinfect the wastewater collected from the STPs. In order to find the least concentration of sodium hypochlorite that result in identifying maximum number of RNA copies using RT-PCR, we performed an optimisation step. We collected 1 L of wastewater in the presence of 20 mL of six different initial concentrations of sodium hypochlorite (Table 1) from the 10 MLD STP. Among the tested concentrations, samples collected with 20 ml of 0.1% hypochlorite resulted in better detection of SARS-CoV-2 RNA. It was also observed that the final concentration of ≥0.04% (initial concentration of ≥ 2%) did not yield any result for the same set of samples, indicating a possible complete inactivation of viral materials by hypo (Table 1; Figure 2). Hence, we used 20 mL of 0.1% sodium hypochlorite per litre of wastewater for all further sample collections.

**Figure 2:**
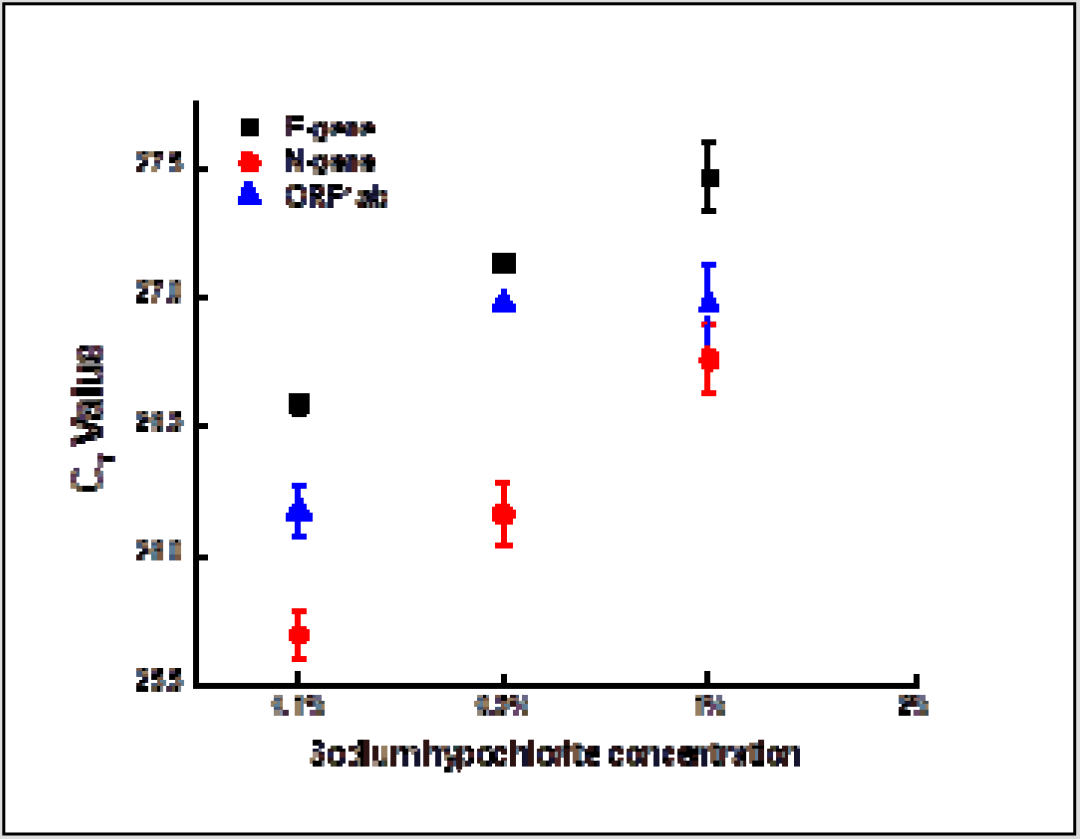
Concentration of sodium hypochlorite affects the detection ability of SARSCoV-2 RNA. Scatter plot showing the effect of different concentrations of sodium hypochlorite on the C_T_ values of viral targets. Each dot represents average C_T_ values obtained from two replicates and the bar represents the standard error of mean.

### 3.2 Detection of SARS-CoV-2 in various STPs and residential community

RT-PCR based detection of SARS-CoV-2 RNA was used for screening the inlet water in the Sewage Treatment Plants (STPs) that cover about 80% of Hyderabad’s STP capacity i.e. 603.5 MLD. SARS-CoV-2 RNA was detected in the inlets of all the tested STPs (Table 2), indicating that the infection is widespread. We observed that the level of viral RNA in the STPs was dynamic, as implied by the changes in C_T_ values of the samples collected on different days (Table 2). We also surveyed samples collected from a gated residential community where confirmed positive cases were reported during the sample collection period and observed the presence of SARS-CoV-2 RNA in the samples in trace amounts (Table 3). As a testimony of efficient wastewater treatment, no viral RNA copies were detected in the outlet of the STPs that we sampled (Table 2).

**Table 2:**
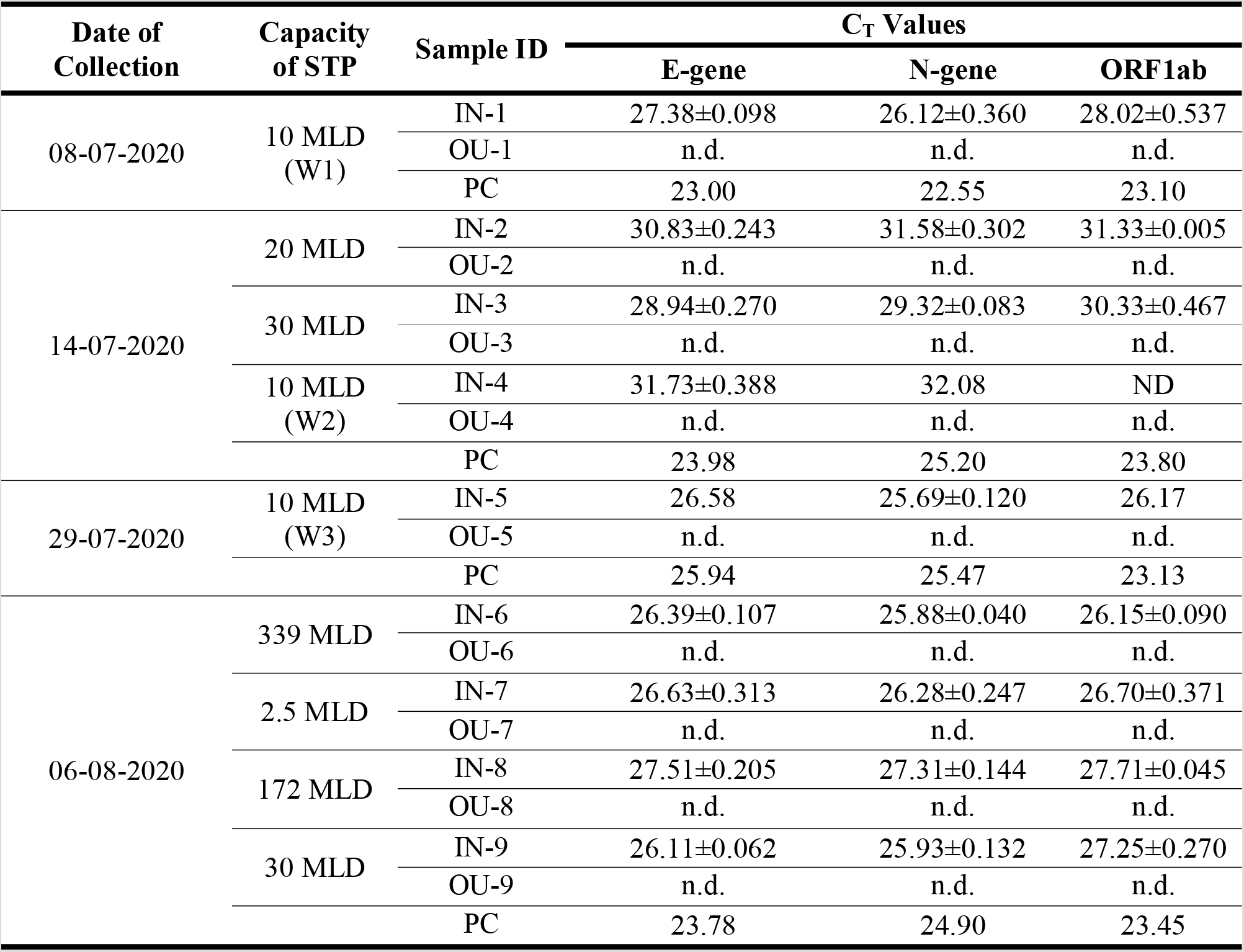
SARS-CoV-2 RNA C_T_ values detected by real-time RT-PCR of raw sewage samples collected from various STP of Hyderabad Metropolitan City. IN = inlet; OU = outlet; W = week; n.d. = not detected; PC = positive control.

**Table 3:**
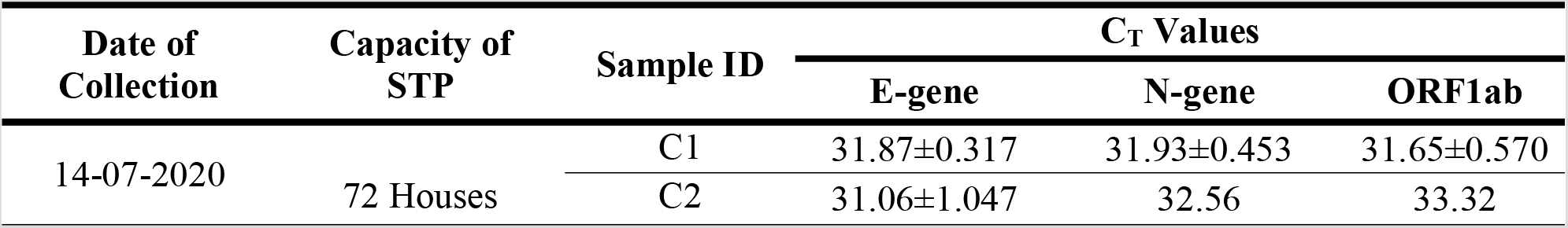
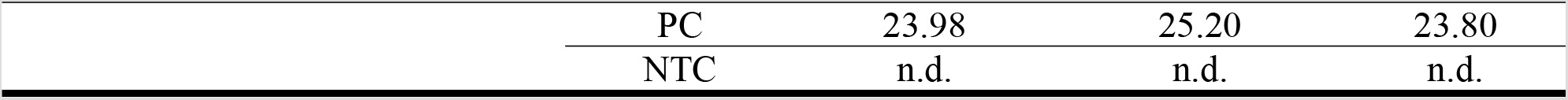
SARS-CoV-2 RNA C_T_ values detected by real-time RT-PCR of raw sewage samples of selected gated community. n.d. = not detected; PC = positive control; NTC = no template control. IN = inlet; OU = outlet; W = week; n.d. = not detected; PC = positive control; NTC = no template control.

### 3.3 Long Term viral load monitoring

One of the STPs (10 MLD), was sampled at different time to assess the dynamics of disease spread with time. We observed a highly fluctuating pattern of viral RNA presence with time, from as low as 30818 copies per L wastewater to as high as 266360 copies per L wastewater (Table 4). The reason for variations could be sampling time, number of actually infected people and the amount of viral shedding by infected individuals, and temporal presence of other compounds (such as surfactants) that could affect the viral material stability in the domestic sewage.

**Table 4:** SARS-CoV-2 RNA C_T_ values detected by real-time RT-PCR of raw sewage samples of selected STP (10 MLD) for weekly monitoring.

| Date of Collection | Week | Sample ID | C<sub>T</sub> Values |  |  | Average number of RNA copies/L water |
| --- | --- | --- | --- | --- | --- | --- |
|  |  |  | E-gene | N-gene | ORF1ab |  |
| 08-07-2020 | W1 | IN-1 | 27.38±0.098 | 26.12±0.360 | 28.02±0.537 | 219540 |
|  |  | OU-1 | n.d. | n.d. | n.d. |  |
|  |  | PC | 23.00 | 22.55 | 23.10 |  |
| 14-07-2020 | W2 | IN-4 | 31.73±0.388 | 32.08 | n.d. | 30818 |
|  |  | OU-4 | n.d. | n.d. | n.d. |  |
|  |  | PC | 23.98 | 25.20 | 23.80 |  |
| 29-07-2020 | W3 | IN-5 | 26.58 | 25.69±0.120 | 26.17 | 266360 |
|  |  | OU-5 | n.d. | n.d. | n.d. |  |
|  |  | PC | 25.94 | 25.47 | 23.13 |  |

### 3.5. Calculation of number of infected people

We used two previously published methods for calculating the number of infected people from the number of RNA copies in the wastewater samples (Ahmed et al., 2020, Hellmér et al., 2014). These methods take into account the number of RNA copies present in the wastewater and the number of RNA copies present in the faecal matter of infected individuals (Section 2.6). Previous studies have established these numbers and we used them for calculating the number of infected individuals (Table 5). Our data indicates that approximately 6.57% of Hyderabad’s population is infected during the window of our study period (Table 6). Existing reports suggest that an infected person shed viral material in faeces for up to 47 days since the symptom onset and remains infectious till 14 days since symptom onset (Wu et al., 2020; Foladori et al., 2020). This suggests that for approximately 35 days, a person sheds viral material while not being infectious. This indicates that 2 in 5 infected people are infectious at any given point of time during the 35 days window. We used this fact to calculate the number of infected people in the active phase of infection (Table 5). Our calculation indicates that approximately 2.6% of the total population of Hyderabad are potentially infectious during the study period.

**Table 5:**
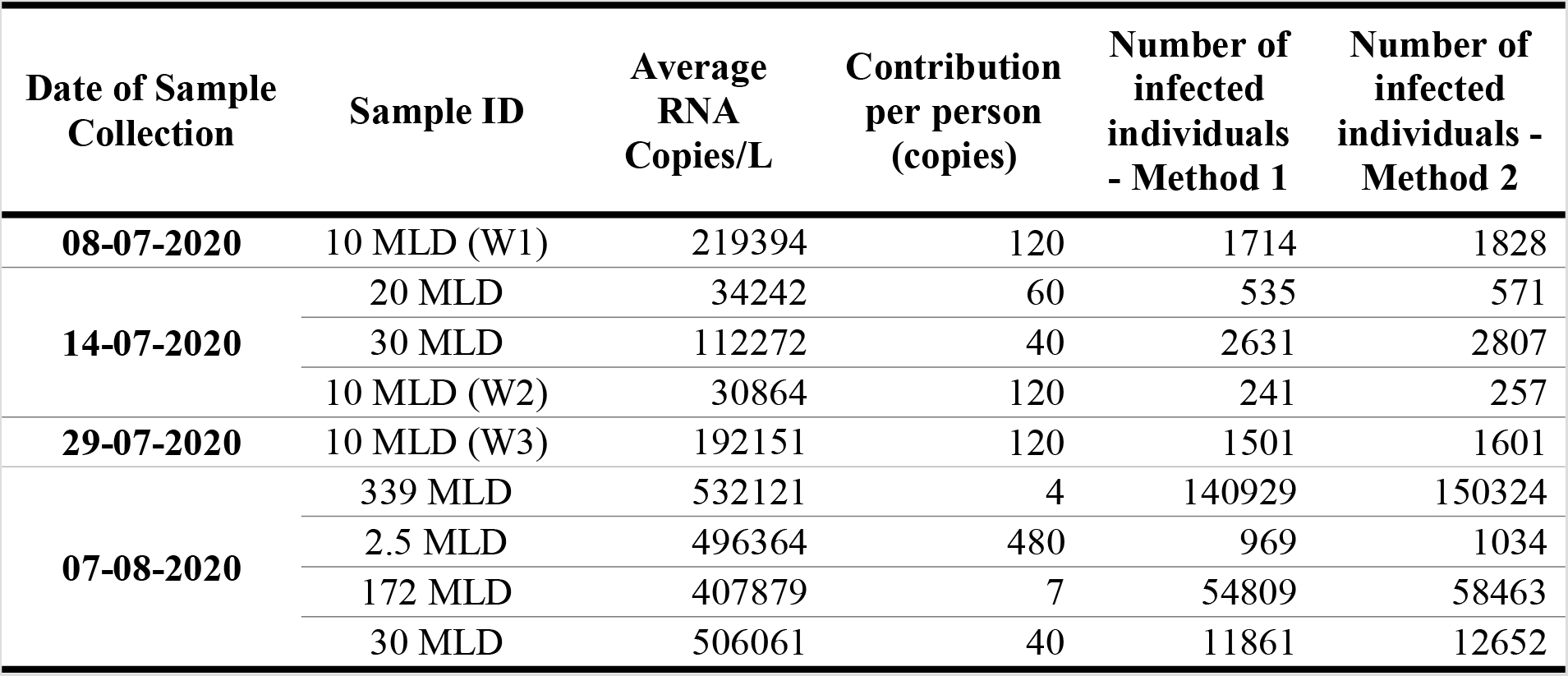
Estimated number of people infected in the areas surveyed through WBE.

**Table 6:** Extrapolation of our data shows the total number of infected and potentially active people in the city of Hyderabad.

|  |  |
| --- | --- |
| Capacity of STP covered (MLD) | 603.5 |
| Average no. of infected people during the window (07-07-2020 to 08-08-2020) | 220343 |
| Total STP capacity of Hyderabad (MLD) | 772 |
| Number of infected (extrapolating to total STP capacity) | 281864 |
| Total wastewater generated in Hyderabad (MLD) | 1800 |
| Number of infected (extrapolating to entire population) | 657197 |
| Population of Hyderabad | 10004144 |
| % Virus shedding individuals | 6.57% |
| Fraction of active cases in the population | 0.4 |
| Number of individuals in potentially active phase of infection | 262878 |

## 4. Conclusion

Our study provides a concrete evidence for the application of WBE as a potential method for disease surveillance. Here we performed an extensive survey of the STPs in Hyderabad for the presence of SARS-CoV-2 genome traces in wastewater. As a result, we observed the presence of SARS-CoV-2 genetic material in all the studied STPs and calculated the extent of spread by estimating the number of infected people and possible number of active cases. These results will be of immense resource for the healthcare and associated departments to vigilantly allocate the necessary resources to manage existing cases as well as to carefully contain the disease spread. Hence, sewage-based surveillance provides a holistic approach to manage the pandemic and also to monitor for future outbreaks, if any. The current study also offers a framework to monitor other pathogens which may be useful managing future epidemics.

## Data Availability

Data referred is available on request.

## Author’s contribution

RKM, SVM and SKK conceptualized the idea. MH, UK, KH and CGG performed the experiments. MH, KH, SVM, UK, SKK, and CGG wrote the manuscript. All the authors have read and approved the manuscript.

## Declaration

The authors declare no conflict of interest.

## Acknowledgments

We acknowledge the Officials of Hyderabad Metropolitan Water Supply and Sewerage Board (HMWSSB) and Hyderabad Metropolitan Development Authority (HMDA), Government of Telangana for providing the samples for this study. UK thanks UGC, CGG and MH thank CSIR for the financial support received. MH, KH, SVM acknowledge the Director, CSIR-IICT for the support. All the authors acknowledge the support received from CSIR, India.

---

**Supplementary Table 1:**
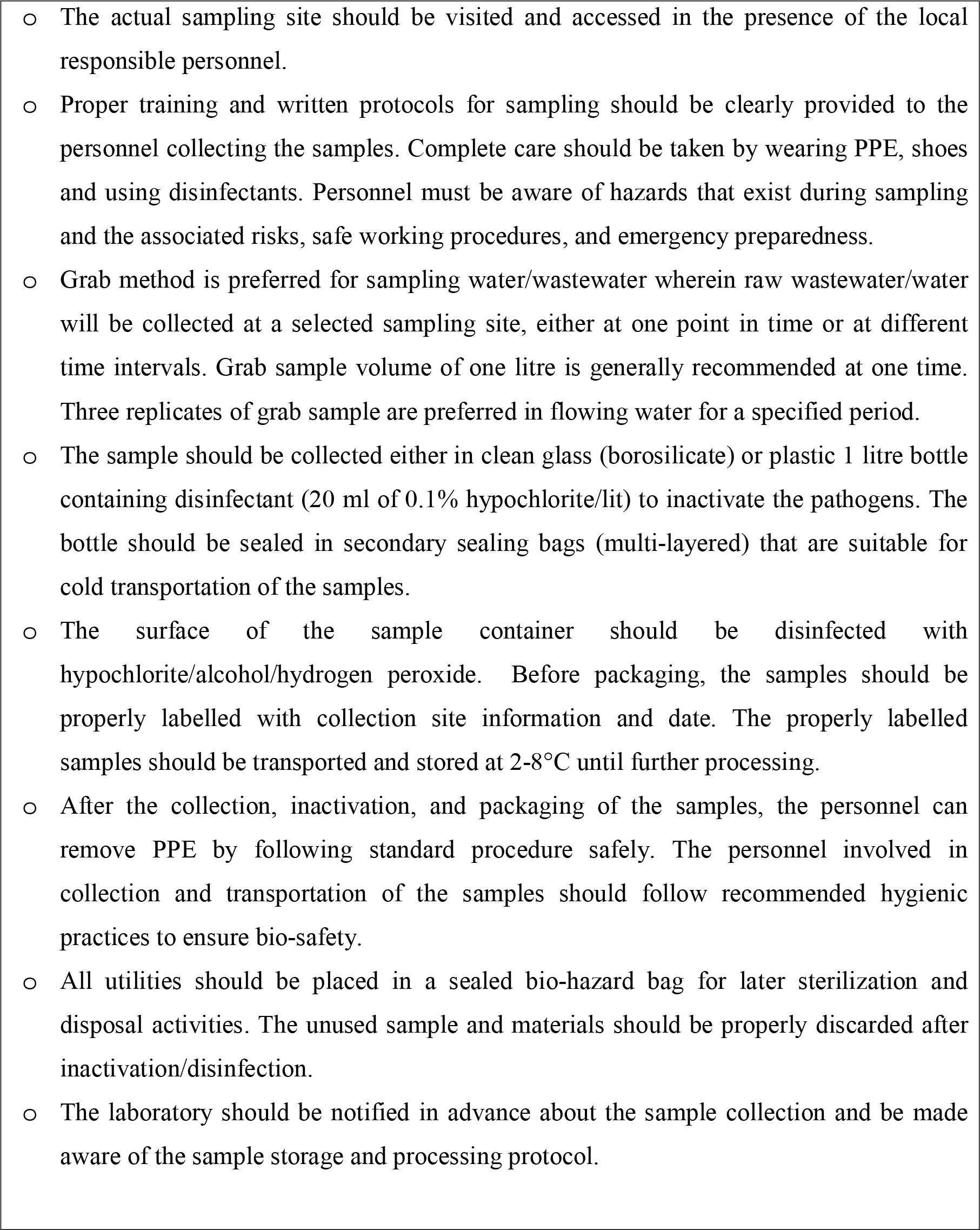
Standard Operating Procedure (SOP) for sampling of wastewater for the detection of viral genome and wastewater-based epidemiological studies

## References

1 W. J. McKibbin, R. Fernando. “The global macroeconomic impacts of COVID-19.” Brookings Institute, no. March (2020): 1–43.

2 W. Wang, Y. Xu, R. Gao, R. Lu, K. Han, G. Wu, W. Tan, Detection of SARS-CoV-2 in different types of clinical specimens. Jama, 323(18) (2020), 1843–1844.

3 F. Xiao, M. Tang, X. Zheng, Y. Liu, X. Li, H. Shan, Evidence for gastrointestinal infection of SARS-CoV-2. Gastroenterology, 158(6) (2020), 1831–1833.

4 H. Zhang, Z. Kang, H. Gong, D. Xu, J. Wang, Z. Li, X. Cui, J. Xiao, T. Meng, W. Zhou, J. Liu, The digestive system is a potential route of 2019-nCov infection: a bioinformatics analysis based on single-cell transcriptomes. BioRxiv. (2020)

5 B.E. Young, S.W.X. Ong, S. Kalimuddin, J.G. Low, S.Y. Tan, J. Loh, O.T. Ng, K. Marimuthu, L.W. Ang, T.M. Mak, S.K. Lau, Epidemiologic features and clinical course of patients infected with SARS-CoV-2 in Singapore. Jama, 323(15) (2020), 1488–1494.

6 R. Woelfel, V.M. Corman, W. Guggemos, M. Seilmaier, S. Zange, M.A. Mueller, D. Niemeyer, P. Vollmar, C. Rothe, M. Hoelscher, T. Bleicker, Clinical presentation and virological assessment of hospitalized cases of coronavirus disease 2019 in a travel-associated transmission cluster. MedRxiv. (2020).

7 M.L. Holshue, C. DeBolt, S. Lindquist, K.H. Lofy, J. Wiesman, H. Bruce, C. Spitters, K. Ericson, S. Wilkerson, A. Tural, G. Diaz, First case of 2019 novel coronavirus in the United States. N. Engl. J. Med. 382 (2020), 929–936.

8 M. Kitajima, W. Ahmed, K. Bibby, A. Carducci, C.P. Gerba, K.A. Hamilton, E. Haramoto, J.B. Rose, SARS-CoV-2 in wastewater: State of the knowledge and research needs. Sci. Total Environ. (2020), 139076.

9 J. Cai, J. Xu, D. Lin, L. Xu, Z. Qu, Y. Zhang, H. Zhang, R. Jia, X. Wang, Y. Ge, A. Xia, A Case Series of children with 2019 novel coronavirus infection: clinical and epidemiological features. Clin. Infect. Dis. (2020).

10 Y. Ling, S.B. Xu, Y.X. Lin, D. Tian, Z.Q. Zhu, F.H. Dai, F. Wu, Z.G. Song, W. Huang, J. Chen, B.J. Hu, Persistence and clearance of viral RNA in 2019 novel coronavirus disease rehabilitation patients. Chin. Med. J. (2020).

11 Y. Wu, C. Guo, L. Tang, Z. Hong, J. Zhou, X. Dong, H. Yin, Q. Xiao, Y. Tang, X. Qu, L. Kuang, Prolonged presence of SARS-CoV-2 viral RNA in faecal samples. Lancet Gastroenterol. Hepatol. 5(5) (2020), 434–435.

12 G. La Rosa, L. Bonadonna, L. Lucentini, S. Kenmoe, E. Suffredini, Coronavirus in water environments: Occurrence, persistence and concentration methods-A scoping review. Water Res. (2020a), 115899.

13 W. Ahmed, N. Angel, J. Edson, K. Bibby, A. Bivins, J.W. O’Brien, P.M. Choi, M. Kitajima, S.L. Simpson, J. Li, B. Tscharke, First confirmed detection of SARS-CoV-2 in untreated wastewater in Australia: A proof of concept for the wastewater surveillance of COVID-19 in the community. Sci. Total Environ. (2020), 138764.

14 S. Wurtzer, V. Marechal, J.M. Mouchel, Y. Maday, R. Teyssou, E. Richard, J.L. Almayrac, L. Moulin, Evaluation of lockdown impact on SARS-CoV-2 dynamics through viral genome quantification in Paris wastewaters. medRxiv. (2020)

15 G. La Rosa, M. Iaconelli, P. Mancini, G.B. Ferraro, C. Veneri, L. Bonadonna, L. Lucentini, E. Suffredini, First detection of SARS-CoV-2 in untreated wastewaters in Italy. Sci. Total Environ. (2020b) 139652.

16 G. Medema, L. Heijnen, G. Elsinga, R. Italiaander, A. Brouwer, Presence of SARS-Coronavirus-2 in sewage. MedRxiv. (2020).

17 W.J. Lodder, A.M. Buisman, S.A. Rutjes, J.C. Heijne, P.F. Teunis, A.M. de Roda Husman, Feasibility of quantitative environmental surveillance in poliovirus eradication strategies. Appl. Environ. Microbiol., 78(11) (2012), 3800–3805.

18 W.J. Lodder, S.A. Rutjes, K. Takumi, A.M. de Roda Husman, Aichi virus in sewage and surface water, The Netherlands. Emerg. Infect. Dis. 19(8) (2013), 1222.

19 P.I. Lee, P.R. Hsueh, Emerging threats from zoonotic corona viruses-from SARS and MERS to 2019-nCoV. J. Microbiol. Immunol. Infect. (2020).

20 R.S. Quilliam, M. Weidmann, V. Moresco, H. Purshouse, Z. O’Hara, D.M. Oliver, COVID-19: The environmental implications of shedding SARS-CoV-2 in human faeces. Environ. Int. (2020).

21 K.Q. Kam, C.F. Yung, L. Cui, A well infant with coronavirus Disease 2019 (COVID-19) with high viral load. [e-pub ahead of print]. Clin Infect Dis. doi, 10.

22 A. Tang, Z.D. Tong, H.L. Wang, Y.X. Dai, K.F. Li, J.N. Liu, W.J. Wu, C. Yuan, M.L. Yu, P. Li, J.B. Yan, Detection of novel coronavirus by RT-PCR in stool specimen from asymptomatic child, China. Emerg. Infect. Dis., 26(6) (2020), 1337–1339.

23 W. Lodder, A.M. de Roda Husman, SARS-CoV-2 in wastewater: potential health risk, but also data source. Lancet Gastroenterol. Hepatol. 5(6) (2020), 533–534.

24 S. Mallapaty, How sewage could reveal true scale of coronavirus outbreak. Nature, 580(7802) (2020), 176–177.

25 C. G. Daughton, “Monitoring Wastewater for Assessing Community Health: Sewage Chemical-Information Mining (SCIM).” Sci. Total Environ. (2018) 619–620 748–764.

26 K. Mao, H. Zhang, and Z. Yang, Can a paper-based device trace COVID-19 sources with wastewater-based epidemiology? (2020).

27 https://www.fda.gov/media/137120/download

28 C. Rose, A. Parker, B. Jefferson, E. Cartmell, The characterization of feces and urine: a review of the literature to inform advanced treatment technology. Critical Reviews in Environ. Sci. Technol. 45(17), (2015), 1827–1879.

29 P. Foladori, F. Cutrupi, N. Segata, S. Manara, F. Pinto, F. Malpei, L. Bruni, G. La Rosa, SARS-CoV-2 from faeces to wastewater treatment: What do we know? A review. Sci Total Environ. (2020) 140444

30 M. Hellmer, N. Paxéus, L. Magnius, L. Enache, B. Arnholm, A. Johansson, T. Bergström, H. Norder, Detection of pathogenic viruses in sewage provided early warnings of hepatitis A virus and norovirus outbreaks. Appl. Environ. Microbiol. 80(21) (2014), 6771–6781.

